# “The Role of Maternal Mental Health in Early Development: Evidence from High Altitude Peru”

**DOI:** 10.64898/2026.07.30.26359334

**Authors:** Milagros Alvarado, Dana Charles McCoy, Daniel Mäusezahl, Lena Jäggi, Kristen Hinckley, Maria Catalina Gastiaburu Cabello, Sarah Farnsworth Hatch, Stella Hartinger, Leonel Aguilar, Andreana Castellanos, Günther Fink

## Abstract

Maternal depressive symptoms are a major public health concern in low- and middle-income countries (LMICs), yet evidence on their relationship with early childhood development (ECD) remains mixed. This study examined the association between maternal depressive symptoms and ECD outcomes during infancy and toddlerhood, and whether household composition moderated these associations. Additionally, we examined whether child stimulation activities were associated with ECD outcomes across both timepoints.

We analysed data from 769 mother-child dyads drawn from a cluster-randomised controlled trial conducted in the Cajamarca region of Peru. Maternal depressive symptoms were assessed during infancy (approximately 5 months) using the DASS-21 depression subscale. Child development was assessed during infancy (mean age 5.2 months) using the CREDI and during toddlerhood (mean age 28.7 months) using the GSED Long Form. Associations were examined using linear regression models, adjusting for demographic, household, social, and caregiving covariates. Effect modification by number of siblings under 6 years and number of adults in the household was assessed.

Approximately one in six mothers (17.4%) reported any depressive symptoms. Maternal depressive symptoms were neither associated with child development outcomes in infancy (adjusted β = 0.01, 95% CI [-0.01, 0.01]) nor in toddlerhood (adjusted β = -0.01, 95% CI [-0.07, 0.05]). No significant moderation was observed for either household composition variable. Among the covariates examined in the adjusted models, child stimulation activities showed the strongest independent association with child development at both timepoints, regardless of maternal depressive symptom status.

We found no evidence that maternal depressive symptoms were associated with child development during infancy or toddlerhood, and household composition did not moderate these associations. These findings suggest that the relationship between maternal depressive symptoms and early child development may be more complex than previously assumed in this rural Andean context. However, the absence of an association in this study should not be interpreted as evidence that maternal depressive symptoms are unimportant, as previous research has documented important effects on maternal wellbeing, family functioning, and child outcomes.

## 1. Introduction

Depression is a leading cause of disability worldwide (1), particularly among women during their childbearing years (2). Numerous studies have found that depression is negatively associated with other facets of women’s well-being (e.g., quality of life, social functioning, and physical health) as well as their children’s health and development (2, 3). These effects are especially pronounced during the perinatal and early parenting periods, when women face unique physiological and psychosocial challenges (4). Mothers experiencing poor mental health may be less able to provide the sensitive, responsive caregiving required for optimal infant development, placing her child at increased risk for adverse outcomes (5). However, the extent to which maternal depressive symptoms translate into adverse child outcomes may depend on the caregiving context in which they occur (6).

Much of the evidence linking maternal mental health and child development comes from high-income contexts, where caregiving is predominantly dyadic, centred on the mother-child relationship. In contrast, in many low- and-middle-income countries (LMICs), caregiving responsibilities are distributed among multiple family members, and community networks play a more central role in child-rearing (7). Whether maternal depressive symptoms affect child development through the same caregiving pathways in these culturally distinct settings remains poorly understood (8).

### Maternal Mental Health and Early Caregiving

Infants depend on their caregivers for nutrition, care, comfort, and social interaction. When caregiver-infant interactions are disrupted, early neurological, emotional, and cognitive development can be compromised (9, 10). Such disruptions matter particularly in the earliest years of life, when responsive parent-child relationships and parental support for learning are crucial for promoting early child development (ECD) (11). Maternal depression is one of the most consistent sources of such disruptions, as symptoms of depression, such as anhedonia, sadness, or lack of energy are likely to undermine sensitive and contingent caregiving (12–14). This caregiving disruptions are in turn linked to poorer cognitive, behavioural, and socio-emotional outcomes in early childhood (15). Evidence from large population samples illustrates this further: children exposed to mothers experiencing psychological distress, including symptoms of depression, anxiety, or stress in the first year after birth, showed poorer socio-emotional and behavioural regulation in early childhood, even among mothers without clinical diagnosis (16, 17).

Evidence from LMICs generally suggests a negative association between maternal depression and child development (18, 19). However, the magnitude of these associations appears to vary across contexts, suggesting that other factors may shape the relationship between maternal mental health and child outcomes. In many LMIC settings, caregiving responsibilities are shared among multiple household members (20). These extended family networks, common in rural Andean communities, may buffer children from the effects of maternal distress. Yet whether and how these caregiving environments shape the impact of maternal mental health on child development remains poorly understood, particularly in rural Andean contexts where extended family structures are the norm.

### Factors that Shape Child Development

A growing body of developmental research suggests that children’s outcomes are shaped not only by individual-level risk factors, such as maternal depression, but also by the broader caregiving environment (21, 22). Child development unfolds through dynamic interactions between the child and multiple layers of environmental influence, including family, community and broader sociocultural contexts (23). Therefore, the adverse effects of early exposure to risk factors, such as maternal depression, may be diminished when families have access to other sufficient contextual resources. Protective factors such as emotional support and the presence of other responsive caregivers can buffer the negative effects of maternal depression on children’s developmental outcomes (9, 24). These protective influences are not static; they evolve as children grow and as family structures adapt to economic and social constraints. In LMICs, extended kin networks and communal caregiving arrangements may serve precisely this buffering function, providing children with additional stable relationships and shared caregiving even under conditions of dyadic-attachment disruptions, economic hardship, and limited access to services (25).

Social support is one factor frequently found to reduce the adverse effects of maternal depressive symptoms on child outcomes (26). Support from partners, family members, or the broader community can improve maternal functioning and promote more responsive caregiving (27). Social support can take many forms, including emotional support, practical help with household tasks, or childcare assistance, and may be especially critical in the postpartum period when mothers are adjusting to new caregiving demands (28, 29). Higher levels of perceived support have been associated with lower rates of maternal depression, higher maternal sensitivity, and more positive developmental outcomes in children (30).

Frequent engagement in stimulation activities such as singing, reading, or playing has been linked to improved cognitive and socio-emotional development, even among children exposed to maternal depression (11, 31). These activities foster early language acquisition, executive function, and emotion regulation, all of which are foundational for later learning and adjustment (32). Stimulation practices may be sensitive to the caregiver’s mental health status, though the extent to which maternal depressive symptoms disrupt these interactions may vary depending on symptom severity and the broader caregiving context. Even in the context of maternal depression, structured opportunities for play and learning may offer children consistent experiences that support resilience (33). However, because the caregiving environment often extends beyond the mother-child relationship, household composition may also shape children’s developmental opportunities.

Household composition (e.g., the presence of other adults or siblings) may also influence caregiving quality in multiple ways. These influences can operate in opposite directions depending on family dynamics and resource availability (20, 34). Larger households can be a source of both support and strain. For example, additional adults might contribute to child supervision or emotional support for the mother, but they can also create crowding or economic pressure (35). The presence of young siblings may compete for the caregiver’s time and energy, which may amplify the effect of maternal depressive symptoms on child development especially in contexts of limited material or emotional resources (36). These dual effects suggest that household composition may either buffer or exacerbate the association between maternal depressive symptoms and child outcomes. Support, however, is not limited to what happens within the household, as families are also embedded in wider social networks that shape maternal wellbeing and child development.

Community-level factors, including participation in social assistance programmes have also been associated with lower maternal mental health distress and better child health outcomes in LMICs (37–39). Whether these associations hold in culturally distinct contexts, such as rural Andean communities, remains understudied.

Together, these factors represent key aspects of the caregiving environment that may shape whether the effects of maternal mental health problems on child development are amplified or mitigated. Their interplay may look particularly different in contexts where caregiving is distributed, resources are constrained, and cultural norms shape how distress is experienced and expressed.

### The Peruvian Andean Context

The Andean region of Peru represents an interesting context to study family-child dynamics. Andean communities are characterised by distinct geographic barriers, strong cultural traditions, close family networks, and persistent socioeconomic challenges. Although Peru has experienced notable economic growth over recent decades, this progress has been unevenly distributed. Rural highland populations continue to face persistent structural challenges, including high levels of poverty, food insecurity, and limited access to health and mental health services (40–43). Cultural stigma around mental illness and traditional gender roles may further discourage women from seeking help, compounding the burden of untreated maternal mental health conditions (44). Structural barriers do not operate in isolation; they intersect with interpersonal stressors that further shape maternal wellbeing in this region.

High rates of intimate partner violence, gender inequities, and unequal caregiving responsibilities further contribute to maternal mental health burdens in this region (45). As a result, maternal mental health concerns are prevalent and often undertreated. These challenges are particularly visible in Cajamarca, a rural Andean region that exemplifies these structural and interpersonal pressures. The current study takes place within this region, where qualitative evidence also highlights local caregiving strengths that may shape how these burdens are experienced and managed.

Qualitative evidence from Cajamarca highlights local caregiving strengths, such as shared childcare responsibilities and intergenerational caregiving, which may sustain child development even in challenging contexts (46). These caregiving strategies may offer critical forms of emotional and instrumental support that buffer both children and mothers from the full effect of psychosocial stressors. For example, when caregiving duties are distributed among multiple adults, mothers may experience reduced stress and improved well-being, which can in turn support more sensitive interactions with their children. Shared caregiving may also provide children with additional stable attachments relationships and opportunities for social learning, while reinforcing cultural continuity and family cohesion (46, 47). Especially in contexts where access to institutional health and social services is limited, these informal and culturally embedded caregiving practices may serve as key protective mechanisms. Whether these caregiving arrangements moderate the relationship between maternal mental health and child development in this context, and which factors shape child outcomes when that association is absent, remains an open empirical question.

### The Present Study

The present study examines the association between maternal depressive symptoms and early childhood development (ECD) outcomes in the Peruvian Andes. It focuses on two key developmental periods: infancy (approximately 5 months old) and toddlerhood (approximately 28 months). Infancy was selected to capture the period immediately following birth, when maternal depressive symptoms is more likely to shape caregiver-infant interactions and attachment formation. Toddlerhood represents a stage when cognitive, socioemotional, and behavioural skills become consolidated and observable and when the cumulative effects of early caregiving experiences may become better detectable (9). This temporal spacing allows for both concurrent and prospective examination of the associations between maternal depressive symptoms and child development.

Before examining these associations, we describe the prevalence and sociodemographic distribution of maternal depressive symptoms in this sample to characterise the study population. The study has four objectives. First, to examine the concurrent association between maternal depressive symptoms and child development outcomes in infancy. Second, to examine the prospective association between maternal depressive symptoms in infancy and child development in toddlerhood. Third, to investigate whether household composition moderates these associations. Specifically, we hypothesise that the number of young siblings under 6 years in the home amplifies potential risks associated with maternal depressive symptoms and whether the presence of three or more adults in the household buffer these risks. Fourth, to examine whether child stimulation activities were associated with child development outcomes across both timepoints. This study seeks to contribute to a nuanced understanding of how maternal depressive symptoms shape early developmental outcomes in a rural Andean community, and whether household structure shapes children’s vulnerability or resilience in this context.

## 2. Methods

### Study Setting and Participants

The study was conducted in three provinces of the Cajamarca region in the northern highlands of Peru: Cajamarca, Cajabamba, and San Marcos. These provinces encompass urban and rural communities at altitudes ranging from 1900 to 3900 meters above sea level (39). Local economies are largely driven by small-scale agriculture and livestock, along with employment in sectors such as education, healthcare, construction, and mining (48).

The data used in this study are drawn from a recently completed cluster-randomised controlled trial (RCT) evaluating a digital infant stimulation platform (ClinicalTrials.gov: NCT05202106) (49) Hereafter this trial is referred to as the parent trial. The RCT comprised four study arms: a home visit group, an AI-supported digital parenting chatbot group, a control group, and a fourth arm comprising mothers already enrolled in the national early childhood programme Cuna Más, who received no study intervention and served as an additional comparison group. Randomisation was conducted at the community level, with 164 clusters serving as the unit of randomisation, each cluster corresponding to a village in rural areas or a block in urban areas. Clusters were allocated across arms as follows: 70 to the control group, 70 to the digital intervention group, and 20 to the home visit group, stratified by province. Both interventions delivered age-appropriate guidance on early stimulation activities, caregiver-child interaction, and responsive caregiving practices. The content focused on promoting early learning and development and did not include components specifically targeting maternal mental health. No significant intervention effects on maternal depressive symptoms were observed in the parent trial. Eligible participants were mothers of infants between 3 and 9 months of age at enrolment, residing in the study area throughout the trial period, and who provided written consent. Recruitment was conducted from September 2021 to March 2023 through a combination of community outreach and referrals from local health centres.

Given that child development at infancy was assessed at baseline prior to any intervention exposure, intervention arm was not included as a covariate in infancy models. For toddlerhood models, intervention arm was included as a covariate to account for potential differential exposure across study arms, and its potential moderating role was examined as a sensitivity analysis.

### Design and Procedure

Although the data derive from an RCT, the present study uses an observational analytic approach to examine associations between maternal depressive symptoms and child development outcomes. Maternal depressive symptoms were measured at infancy. Child development outcomes were assessed concurrently during infancy and prospectively during toddlerhood using validated instruments described in the Measures section. Analysis was restricted to dyads with complete data on both exposure and outcome variables (complete-case analysis).

**Figure 1.**
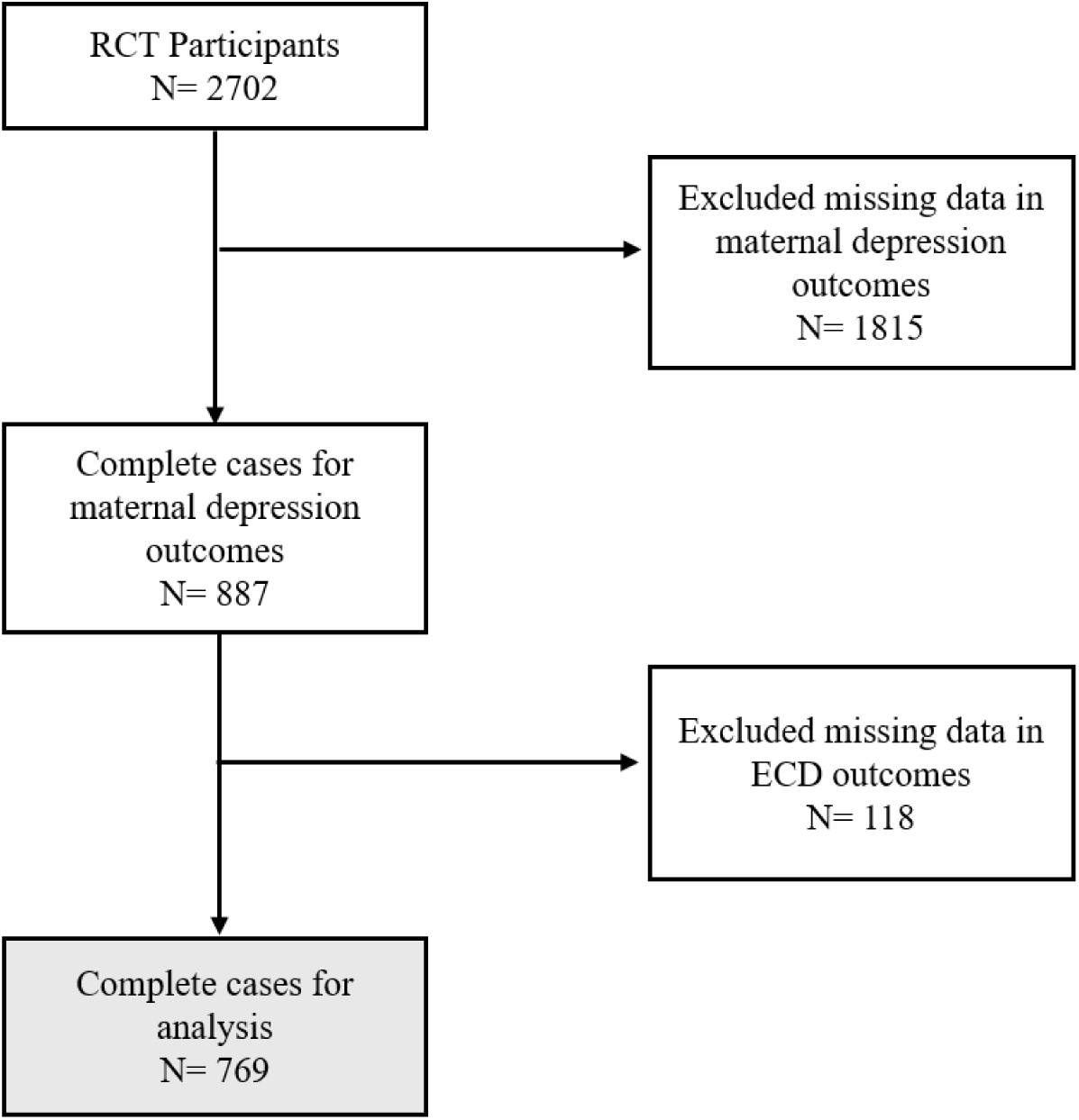
Flowchart of mother-child dyads included in the analysis.

The sample comprised 769 mother–child dyads with complete data on maternal depressive symptoms and child developmental outcomes at both timepoints. At infancy, the mean age of children was 5.2 months; at toddlerhood it was 28.7 months. Child sex was approximately evenly distributed, with 48% female. Mothers were predominantly aged between 20 and 30 years, and households had on average 2.5 adult occupants. Full descriptive characteristics of the sample are presented in Table 1. Comparisons between dyads with and without complete data revealed no significant differences in child sex, maternal age, maternal education, place of residence, household overcrowding, or socioeconomic status. Child age and intervention arm differed between groups and were therefore included as covariates in all toddlerhood models.

**Table 1.**
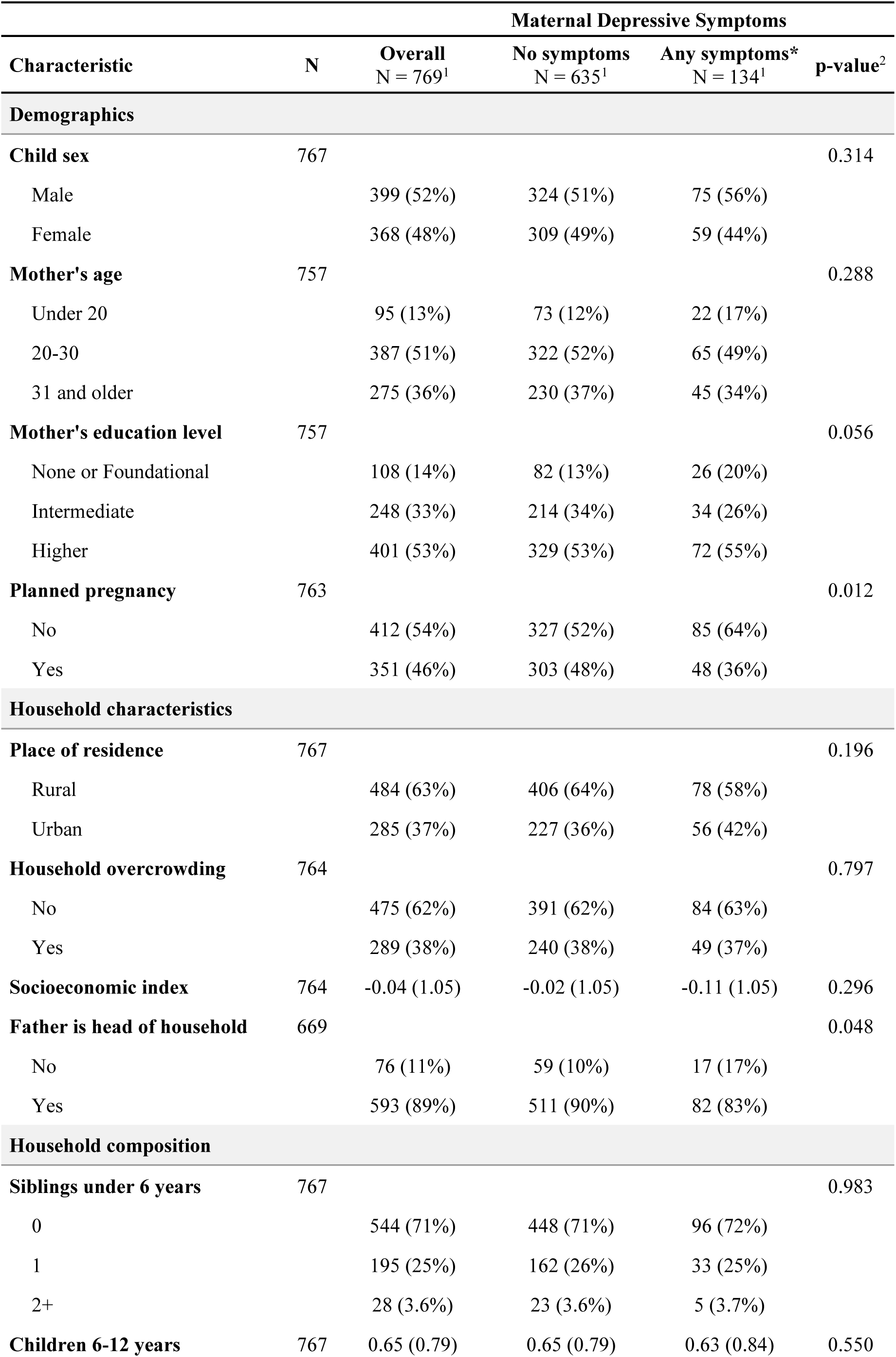
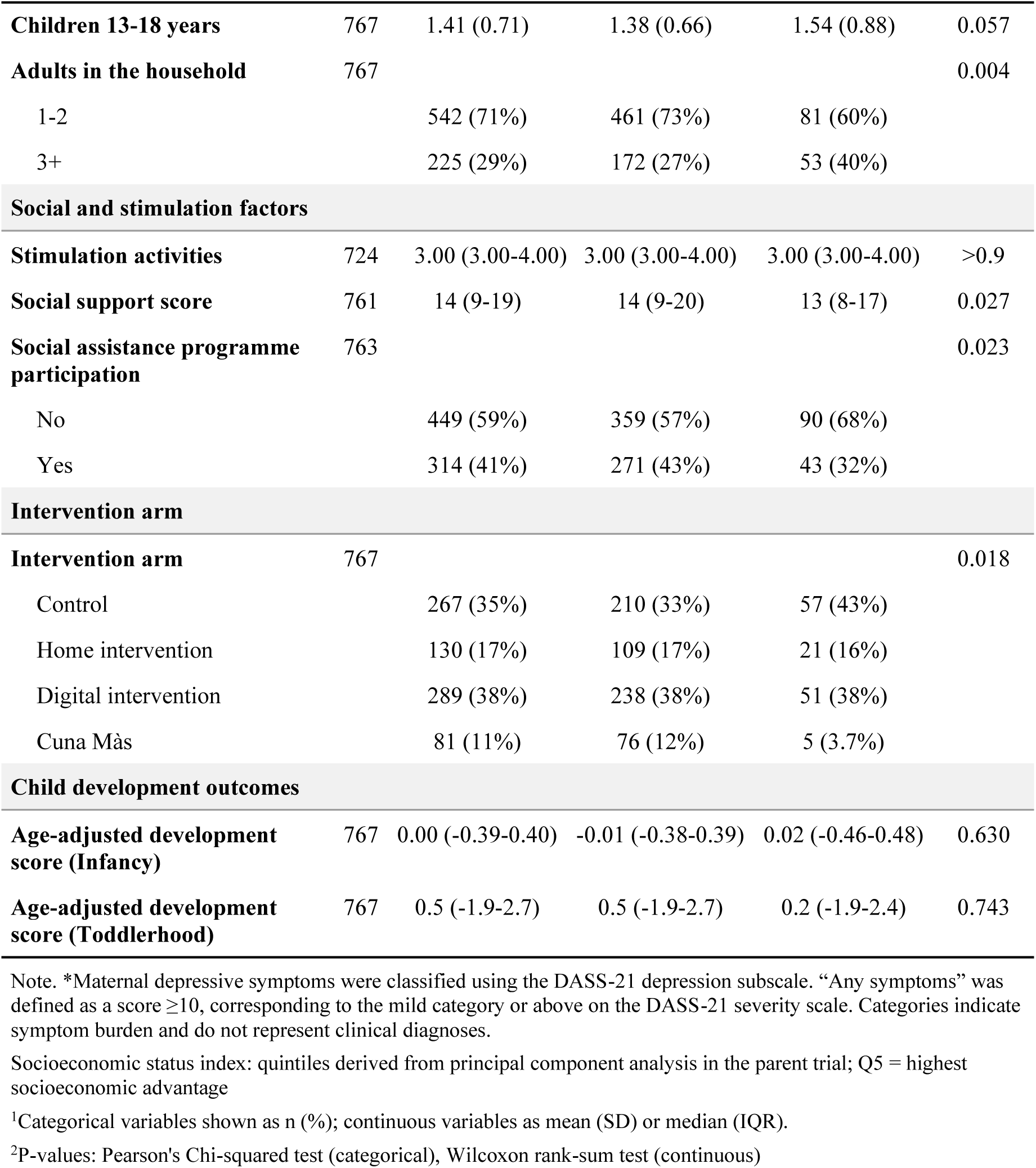
Baseline characteristics by maternal depressive symptom status in infancy.

The research protocol was reviewed and approved by the relevant institutional ethics review boards prior to the initiation of participant recruitment: Universidad Peruana Cayetano Heredia (SIDISI: 214395) and Ethikkommission Nordwest- und Zentralschweiz (AO 2024-00100). All families provided written consent prior to their participation in the study.

### Measures

#### Maternal Depressive Symptoms

Maternal depressive symptoms were assessed using the depression subscale of the DASS-21, a validated self-report instrument administered by trained interviewers who read each item aloud to participants. Because the DASS-21 measures depressive symptomatology rather than providing a clinical diagnosis, we refer to these scores as maternal depressive symptoms throughout the manuscript. The depression scale includes 7 items (items 3, 5, 10, 13, 16, 17, and 21) assessing symptoms such as hopelessness, low mood, and lack of interest. Items are rated on a 4-point Likert scale ranging from 0 (“Did not apply to me at all”) to 3 (“Applied to me very much or most of the time”), with total subscale scores ranging from 0 to 21. Observed scores in this sample ranged from 0 to 21. The subscale demonstrated high internal consistency in this sample (Cronbach’s α = 0.836, 95% CI [0.818, 0.854]; see Supplementary Materials, Table S1).

In all regression analyses, continuous sum scores were used to estimate associations between maternal depressive symptoms and child development outcomes. This approach preserved the full variability in symptom severity and avoided imposing potentially non-validated categorical thresholds. For descriptive purposes, a dichotomous variable was created to indicate the presence or absence of depressive symptoms (50). A score of ≥10 was used to classify participants as experiencing any level of symptoms, corresponding to the mild category or above on the DASS-21 severity scale (categories: normal, mild, moderate, severe, extremely severe).

#### Child Development Outcomes

In infancy, child development was assessed using the Caregiver Reported Early Development Instrument (CREDI), a caregiver-report tool designed to capture age-appropriate milestones in children under three years (51). The CREDI assesses development across four domains: cognitive, language, motor, and socio-emotional functioning. The CREDI produces overall z-scores normalised within each age group in a global reference sample, with higher scores indicating more advanced development relative to same-age peers. For the present analyses, raw scores were used as the outcome variable in infancy models, with child age in days included as a covariate

In toddlerhood, child development was assessed using the Global Scales for Early Development Long Form (GSED-LF), an open-access tool designed to measure development in children up to 36 months across motor, cognitive, language, and socio-emotional domains (52). Assessment was conducted through direct assessment by trained data collectors. The GSED-LF produces a D-score that is not age-standardised; child age in days at assessment was therefore included as a covariate in all toddlerhood models (52).

#### Household Composition Factors

Household composition variables were examined both as covariates and as potential moderators of the association between maternal depressive symptoms and child development. The number of siblings residing in the home was captured across three age bands, categorised as 0, 1, or 2+ children in each band: children under 6 years, children aged 6-12 years and children aged 13-18 years. The number of adults in the household was categorised as 1-2 versus 3 or more, with the latter reflecting the presence of an extended family network. Of these variables, two were additionally tested as moderators: siblings under 6 years, based on theoretical hypotheses about competing caregiving demand and number of adults in the household, based on hypotheses about extended family support (see Analytic Plan). In moderation models, each variable served as the focal moderator while the other was retained as a covariate.

#### Covariates

Covariates were selected based on theoretical and empirical associations with maternal depressive symptoms and ECD and all covariates were measured at infancy. Demographic characteristics included child sex (0 = female, 1 = male), maternal age (categorised as under 20, 20–30, and 31 or older), maternal education level (coded ordinally as none or foundational, intermediate, and higher), whether the pregnancy was planned (0 = no, 1 = yes), and child age in months or days at the time of developmental assessment.

Household characteristics captured structural and economic dimensions of the home environment, including place of residence (0 = rural, 1 = urban), household overcrowding, defined as the ratio of household members to number of bedrooms (0 = no, 1 = yes), and household socioeconomic status measured using an asset-based index generated by principal components analysis. Quintiles were derived from the first principal component in the parent trial, with higher quintiles indicating greater socioeconomic advantage (53). Paternal presence was captured as a binary variable indicating whether the biological father was identified by the mother as the household head at the time of assessment (0 = no, 1 = yes). Beyond household structure, social and caregiving factors were also considered. These included perceived social support, stimulation practices, and social assistance programme participation. Perceived social support was measured using a composite score of 8 items rated on a 4-point Likert scale (0 = never, 3 = always), with total scores ranging from 0 to 24. Higher scores indicate greater perceived emotional and instrumental support from key individuals (54). Stimulation activities were assessed using six items adapted from the UNICEF Multiple Indicator Cluster Survey (MICS) module, capturing whether the child had received any of the following activities from any household member aged 15 years or older in the three days preceding the survey: jointly reading books or looking at pictures, telling stories, singing song or lullabies, taking the child for a walk, playing with the child, and naming, counting or drawing things with the child. Responses were summed to produce a total score ranging from 0 to 6 with higher scores indicating that the child experienced a greater number of stimulation activities (55). Social assistance programme participation was operationalised as a binary variable indicating whether the primary caregiver participated or any household member had participated in a formal or informal social programme in the previous 12 months (0= no, 1= yes).

### Analytic Plan

The analytic plan followed three sequential steps. First, sample demographic and household characteristics were described overall and by maternal depressive status (no symptoms vs any symptoms) at infancy. To assess potential bias, key demographic and household variables were compared between dyads with and without complete data. Full details of this analysis are provided in the Supplementary Materials (Table S2). Baseline characteristics were broadly balanced across study arms, except for the Cuna Más group, which differed in several sociodemographic characteristics reflecting its targeted enrolment criteria (Supplementary Materials, Table S3). These descriptive analyses informed the selection of covariates for subsequent regression models. As sensitivity analyses, we additionally tested for non-linear associations between maternal depressive symptoms and child development by including quadratic terms in the adjusted models at both timepoints (Supplementary Materials, Table S4) and examined whether intervention arm modified the association between maternal depressive symptoms and child development at toddlerhood (Supplementary Materials, Table S5).

Second, associations between continuous maternal depressive symptom scores and child developmental outcomes were examined using linear regression. Separate models were estimated for each developmental timepoint: CREDI raw-scores at infancy and GSED D-scores at toddlerhood adjusted for child age. For infancy models, an unadjusted model was estimated first, followed by an adjusted model including a prespecified set of covariates selected based on theoretical and empirical associations with maternal depressive symptoms and ECD, covering demographic characteristics, household conditions and composition, and social and stimulation factors. For toddlerhood models, the same covariate structure was applied with the addition of child age in days at assessment and intervention arm to account for potential differential exposure across study arms

Third, effect modification was assessed by adding interaction terms to the adjusted models. Two prespecified moderator variables were examined: number of siblings under 6 years (0, 1, 2+) and number of adults in the household (1–2, 3+). The depression symptom score was mean centred prior to computing interaction terms. Moderation analyses were conducted for both timepoints: infancy and toddlerhood (see Supplementary Materials, Figures S1-S4).

Additionally, associations between stimulation activities and child development outcomes, corresponding to the fourth study objective, were examined through the covariate estimates of the fully adjusted models.

All statistical analyses were conducted in R (*version 4.5.1)*. Regression coefficients are reported as unstandardised estimates with 95% confidence intervals (CIs), reflecting the expected change in child development scores per one-unit increase in maternal depressive symptoms. Unstandardised coefficients were chosen to maintain interpretability in the original measurement units. To facilitate comparison across timepoints, adjusted results are presented together in a forest plot. Stimulation activities were included as a covariate in the fully adjusted models; associations between stimulation and child development outcomes are reported as part of the covariate estimates and were not examined in separate models. To account for the cluster-randomised design, all models were estimated with cluster-robust standard errors using the sandwich estimator (vcovCL in R), clustering observations at the community level.

## 3. Results

### Participant characteristics

The analytic sample included 769 mother-child dyads with complete data on maternal depressive symptoms in infancy and child developmental outcomes at both timepoints. Mothers with any level of depressive symptoms (17.4%, n = 134) were less likely to report that the current pregnancy has been planned (36% vs. 48%, p = 0.012). They also reported lower levels of social support and lower rates of social assistance programme participation. These mothers were also more likely to live in households with three or more adults (40% vs. 27%, p = 0.004). Household overcrowding did not differ between groups. Child age at the time of measurement during toddlerhood was slightly higher among mothers with depressive symptoms (912 vs. 893 days, p < 0.001), which explains the marginally higher GSED D-scores observed in this group, as D-scores increase with age. Study arm distribution differed across groups (p = 0.017), with the control group accounting for 43% of all mothers with depressive symptoms in the sample.

### Prevalence of Maternal Depressive Symptoms in Infancy

Approximately one in six mothers (17.4%, n = 134) reported any depressive symptoms in infancy (DASS-21 score ≥10). The distribution of depression scores was right-skewed, with a mean of 4.94 (SD = 6.47) and a median of 2 (IQR: 0–6), indicating that most mothers reported few or no symptoms whereas a smaller proportion presented with moderate to severe levels.

### Maternal Depressive Symptoms and Child Development

Tables 3 and 4 present unadjusted and adjusted regression estimate for the association between maternal depressive symptoms and child development at infancy and toddlerhood, respectively.

**Table 3.** Estimated Association between Maternal Depressive Symptoms and Child Development in Infancy.

| Characteristic | Unadjusted |  |  | Adjusted |  |  |
| --- | --- | --- | --- | --- | --- | --- |
|  | Beta | 95% CI | p-value | Beta | 95% CI | p-value |
| **Exposure** |  |  |  |  |  |  |
| Maternal depressive symptoms (Infancy) | 0.00 | -0.01, 0.01 | 0.601 | 0.00 | -0.01, 0.01 | 0.753 |
| **Demographics** |  |  |  |  |  |  |
| Child age (months) | 0.72 | 0.68, 0.75 | <0.001 | 0.69 | 0.66, 0.73 | <0.001 |
| Child sex |  |  |  |  |  |  |
| Male |  |  |  | Ref. | - | - |
| Female |  |  |  | -0.01 | -0.12, 0.11 | 0.875 |
| Mother age |  |  |  |  |  |  |
| Under 20 |  |  |  | Ref. | - | - |
| 20-30 |  |  |  | -0.06 | -0.26, 0.14 | 0.566 |
| 31 and older |  |  |  | -0.06 | -0.28, 0.17 | 0.634 |
| Mother education |  |  |  |  |  |  |
| None or foundational |  |  |  | Ref. | - | - |
| Intermediate |  |  |  | 0.03 | -0.14, 0.21 | 0.724 |
| Higher |  |  |  | 0.15 | -0.06, 0.35 | 0.166 |
| **Household characteristics** |  |  |  |  |  |  |
| Place of residence |  |  |  |  |  |  |
| Rural |  |  |  | Ref. | - | - |
| Urban |  |  |  | 0.03 | -0.16, 0.23 | 0.744 |
|  | Beta | 95% CI | p-value | Beta | 95% CI | p-value |
| Socioeconomic index |  |  |  | 0.02 | -0.06, 0.09 | 0.627 |
| **Household composition** |  |  |  |  |  |  |
| Sibling under 6 years |  |  |  |  |  |  |
| 0 |  |  |  | Ref. | - | - |
| 1 |  |  |  | -0.11 | -0.25, 0.02 | 0.094 |
| 2+ |  |  |  | -0.42 | -0.81, -0.02 | **0.040** |
| Children 6-12 years |  |  |  | 0.00 | -0.08, 0.09 | 0.915 |
| Children 13-18 years |  |  |  | 0.02 | -0.06, 0.12 | 0.536 |
| Adults at the household |  |  |  |  |  |  |
| 1-2 |  |  |  | Ref. | - | - |
| 3+ |  |  |  | -0.05 | -0.17, 0.07 | 0.453 |
| **Social and stimulation factors** |  |  |  |  |  |  |
| Stimulation |  |  |  | 0.12 | 0.08, 0.17 | **<0.001** |
| Social support score |  |  |  | 0.01 | 0.00, 0.02 | **0.006** |
| Social assistance programme participation |  |  |  |  |  |  |
| No |  |  |  | Ref. | - | - |
| Yes |  |  |  | -0.14 | -0.29, 0.02 | 0.085 |

Abbreviation: CI = Confidence Interval
*Socioeconomic index – same as above*

**Table 4.**
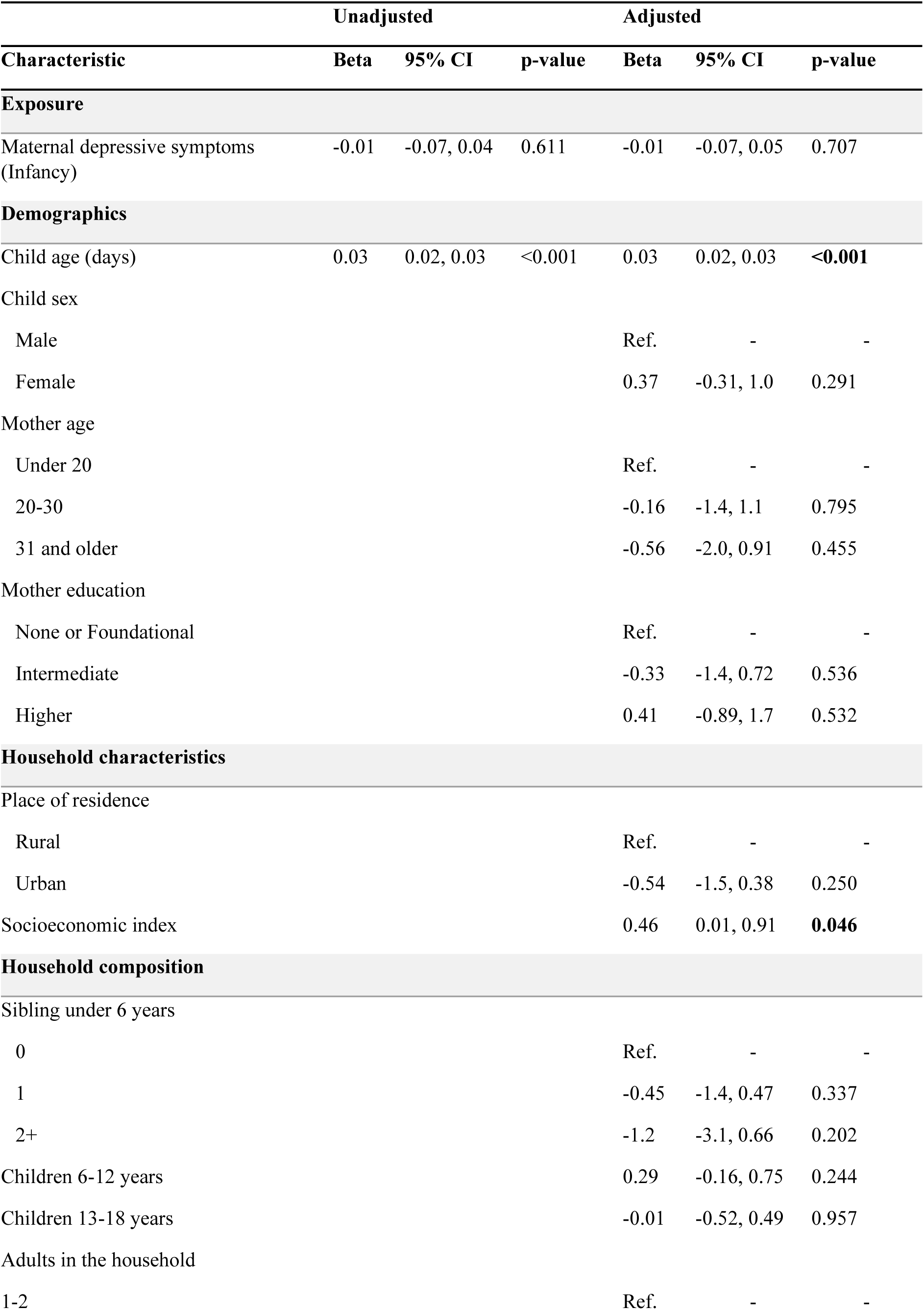
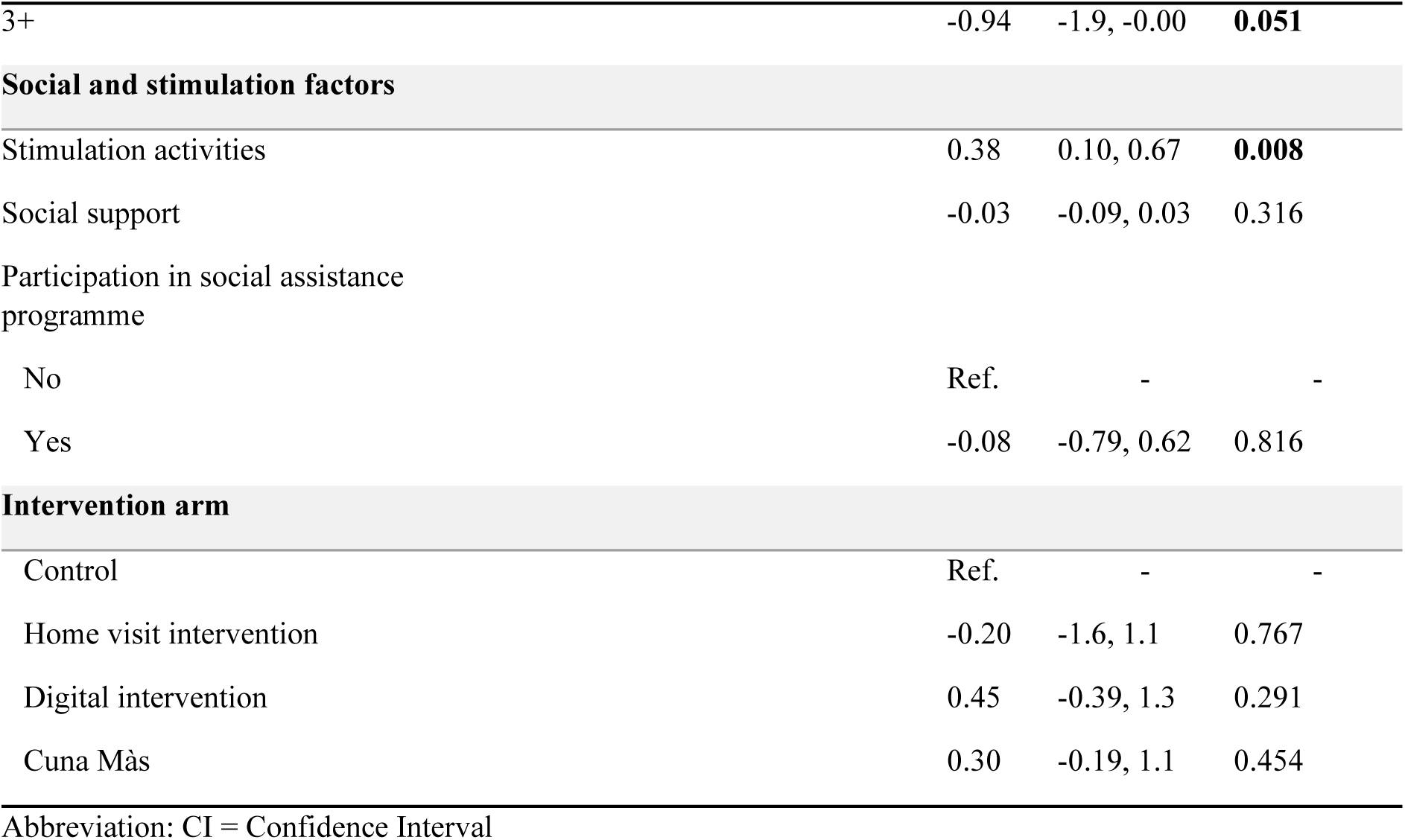
Estimated Association between Maternal Depressive Symptoms and Child Development in Toddlerhood.

| Characteristic | Unadjusted |  |  | Adjusted |  |  |
| --- | --- | --- | --- | --- | --- | --- |
|  | Beta | 95% CI | p-value | Beta | 95% CI | p-value |
| **Exposure** |  |  |  |  |  |  |
| Maternal depressive symptoms (Infancy) | -0.01 | -0.07, 0.04 | 0.611 | -0.01 | -0.07, 0.05 | 0.707 |
| **Demographics** |  |  |  |  |  |  |
| Child age (days) | 0.03 | 0.02, 0.03 | <0.001 | 0.03 | 0.02, 0.03 | **<0.001** |
| Child sex |  |  |  |  |  |  |
| Male |  |  |  | Ref. | - | - |
| Female |  |  |  | 0.37 | -0.31, 1.0 | 0.291 |
| Mother age |  |  |  |  |  |  |
| Under 20 |  |  |  | Ref. | - | - |
| 20-30 |  |  |  | -0.16 | -1.4, 1.1 | 0.795 |
| 31 and older |  |  |  | -0.56 | -2.0, 0.91 | 0.455 |
| Mother education |  |  |  |  |  |  |
| None or Foundational |  |  |  | Ref. | - | - |
| Intermediate |  |  |  | -0.33 | -1.4, 0.72 | 0.536 |
| Higher |  |  |  | 0.41 | -0.89, 1.7 | 0.532 |
| **Household characteristics** |  |  |  |  |  |  |
| Place of residence |  |  |  |  |  |  |
| Rural |  |  |  | Ref. | - | - |
| Urban |  |  |  | -0.54 | -1.5, 0.38 | 0.250 |
| Socioeconomic index |  |  |  | 0.46 | 0.01, 0.91 | **0.046** |
| **Household composition** |  |  |  |  |  |  |
| Sibling under 6 years |  |  |  |  |  |  |
| 0 |  |  |  | Ref. | - | - |
| 1 |  |  |  | -0.45 | -1.4, 0.47 | 0.337 |
| 2+ |  |  |  | -1.2 | -3.1, 0.66 | 0.202 |
| Children 6-12 years |  |  |  | 0.29 | -0.16, 0.75 | 0.244 |
| Children 13-18 years |  |  |  | -0.01 | -0.52, 0.49 | 0.957 |
| Adults in the household |  |  |  |  |  |  |
| 1-2 |  |  |  | Ref. | - | - |
|  | Beta | 95% CI | p-value | Beta | 95% CI | p-value |
| 3+ |  |  |  | -0.94 | -1.9, -0.00 | **0.051** |
| **Social and stimulation factors** |  |  |  |  |  |  |
| Stimulation activities |  |  |  | 0.38 | 0.10, 0.67 | **0.008** |
| Social support |  |  |  | -0.03 | -0.09, 0.03 | 0.316 |
| Participation in social assistance programme |  |  |  |  |  |  |
| No |  |  |  | Ref. | - | - |
| Yes |  |  |  | -0.08 | -0.79, 0.62 | 0.816 |
| **Intervention arm** |  |  |  |  |  |  |
| Control |  |  |  | Ref. | - | - |
| Home visit intervention |  |  |  | -0.20 | -1.6, 1.1 | 0.767 |
| Digital intervention |  |  |  | 0.45 | -0.39, 1.3 | 0.291 |
| Cuna Màs |  |  |  | 0.30 | -0.19, 1.1 | 0.454 |

Abbreviation: CI = Confidence Interval

Maternal depressive symptom scores in infancy were neither significantly associated with child development outcomes in infancy (Table 3) nor with child development outcomes in toddlerhood (Table 4). These null findings were stable across all models. Full model results are reported in Supplementary Materials (Tables S7-S8).

In infancy, among the covariates included in the adjusted model, stimulation activities showed the strongest independent association with child development (β = 0.12, p < 0.001). Social support also showed a positive association with child development (β = 0.01, p = 0.006). In contrast, having two or more children under 6 years in the household was negatively associated with child development (β = - 0.42, p = 0.040). All other covariates were not statistically significant.

The null association between maternal depressive symptoms and child development at toddlerhood was consistent across all six models estimated (β = −0.01, p = 0.707). Among covariates included in the toddlerhood model, stimulation activities again showed the strongest independent association with child development with a notably larger coefficient than observed in infancy (β = 0.38, p = 0.008). All other covariates were not statistically significant.

### Moderating effect of household composition

Tables 5 and 6 present moderation analyses by household composition at infancy and toddlerhood respectively. Interaction plots are provided in Supplementary Materials (Figures S1–S4).

**Table 5.** Infancy: Moderation of the association between maternal depressive symptoms and ECD in infancy by household composition.

| Characteristic | Young siblings under 6 years old |  |  | Adults in household |  |  |
| --- | --- | --- | --- | --- | --- | --- |
|  | Beta | 95% CI | p-value | Beta | 95% CI | p-value |
| Maternal depressive symptoms* × 1 young sibling | 0.01 | -0.01, 0.02 | 0.278 |  |  |  |
| Maternal depressive symptoms* × 2+ young sibling | 0.00 | -0.05, 0.06 | 0.934 |  |  |  |
| Maternal depressive symptoms* × 3+ Adults in the household |  |  |  | 0.00 | -0.02, 0.01 | 0.545 |

Note. \*The DASS-21 depression score was mean-centred before calculating interaction terms. Reference categories are 0 young siblings aged 0–5 years and 1–2 adults in the household.
Abbreviation: CI = Confidence Interval

**Table 6.** Toddlerhood: Moderation of the association between maternal depressive symptoms and ECD in toddlerhood by household composition.

| Characteristic | Young siblings under 6 years old |  |  | Adults in household |  |  |
| --- | --- | --- | --- | --- | --- | --- |
|  | Beta | 95% CI | p-value | Beta | 95% CI | p-value |
| Maternal depressive symptoms* × 1 young sibling | 0.01 | -0.21, 0.22 | 0.958 |  |  |  |
| Maternal depressive symptoms* × 2+ young sibling | 0.07 | -0.08, 0.21 | 0.367 |  |  |  |
| Maternal depressive symptoms* × 3+ Adults in the household |  |  |  | 0.02 | -0.18, 0.14 | 0.780 |

Note. \*The DASS-21 depression score was mean-centred before calculating interaction terms. Reference categories are 0 young siblings aged 0–5 years and 1–2 adults in the household.
Abbreviation: CI = Confidence Interval

Household composition did not moderate the association between maternal depressive symptoms and child developmental outcomes at either timepoint. In infancy, the magnitude of the association did not differ by number of siblings under 6 years in the household, nor by number of adults in the household. Similarly, no moderation was observed at toddlerhood by number of siblings or by number of adults in the household.

## 4. Discussion

This study examined the association between maternal depressive symptoms and early childhood development during infancy and toddlerhood in a rural Andean region of Peru. Remarkably, we did not find any association between maternal depressive symptoms and child developmental outcomes at either timepoint, or household composition did not moderate this association. A notable additional finding is that stimulation activities received by the child showed the strongest independent association with child development at both timepoints, independently of maternal depressive symptom status. This is consistent with evidence linking frequent engagement in stimulation activities to better early cognitive and socio-emotional development in LMIC contexts (32) and suggests that everyday caregiving interactions may be a key correlate of early development in this setting, independent of maternal depressive symptom status (11).

Our findings seem consistent with recent evidence from LMIC settings suggesting that the association between maternal depressive symptoms and child development may be attenuated or absent in contexts where caregiving responsibilities are shared within the household (6). These findings contrast with a substantial body of literature documenting negative associations between maternal depressive symptoms and child cognitive, emotional, and behavioural development (56–59). The Lancet series on Perinatal Mental Health emphasises that parenting quality is a key mediator in this relationship (60), and regardless of the availability of formal mental health support, the presence of informal caregiving networks may play a critical role in buffering children from the effects of maternal depressive symptoms.

A growing body of research suggests that maternal depressive symptoms do not uniformly impair caregiving. In some cases, mothers experiencing mild to moderate depressive symptoms may invest additional effort in childcare to maintain maternal identity or regulate their own emotions, potentially supporting responsive and attentive caregiving (47). This pattern has been observed in high-risk populations, where mothers with depressive symptoms increased time and attention devoted to caregiving, potentially to maintain closeness with their child or manage emotional distress (47). When extended family systems or social networks are available, such compensatory caregiving effort may be further sustained, buffering children from the adverse effects of parental distress. However, such compensatory caregiving effort may be more sustainable under conditions of lower symptom burden, as evidence suggests that more severe and chronic symptoms are associated with greater disruptions to caregiving quality (61).These dynamics may help explain the absence of a negative association between maternal depressive symptoms and child development outcomes commonly reported in other contexts. The characteristics of depressive symptoms themselves, particularly their severity and chronicity, may further shape whether and how they translate into caregiving disruptions and adverse child outcomes.

The strength of associations between maternal depressive symptoms and child development varies depending on symptom characteristics, including severity, timing, and duration (62, 63). Recent evidence suggests that severity and chronicity are each independently associated with child outcomes, with more severe and persistent symptoms showing stronger effects (64). Milder or transient symptoms, consistent with the right-skewed distribution observed in this sample, may therefore have a more limited impact on caregiving quality and child development. Even when symptoms are more severe, their impact may not be uniform across all domains of development (65, 66). Domains that rely heavily on the quality of caregiver–child interactions, such as language and socioemotional development, may be more sensitive to maternal depressive symptoms than domains with a stronger biological basis, such as motor development (67). Depressive symptoms may therefore affect child development less through their mere presence and more through the extent to which they disrupt parenting behaviours, a disruption that may be less pronounced at lower symptom levels. In this sample, stimulation activities received by the child remained strongly associated with child development regardless of maternal depressive symptom status, and stimulation levels did not differ between mothers with and without depressive symptoms (median = 3 in both groups, p > 0.9), suggesting that children’s exposure to stimulating interactions was largely preserved. This pattern may reflect the extent to which everyday caregiving interactions, including stimulation activities, remained intact in this context despite maternal depressive symptoms, whether through mothers’ own continued engagement or through the contribution of other household members. Importantly, our measure captured stimulation received by the child rather than activities performed specifically by the mother, which may itself reflect the distributed caregiving arrangements common in this setting. Understanding who provides these interactions, and under what conditions, remains an important direction for future research (68).

Beyond the parenting behaviours, the broader household caregiving context may also shape whether maternal depressive symptoms translate into developmental consequences for children. Changes in mother-child interaction patterns associated with maternal depressive symptoms do not fully account for child outcomes, highlighting the complex and sometimes indirect pathways through which maternal depressive symptoms shape development (69). In contexts where caregiving is shared across multiple household members, as is common in rural Andean communities, other caregivers may compensate for reduced maternal sensitivity, collectively maintaining the quality of interactions that children need for healthy development. Such collective caregiving arrangements may offer one potential pathway through which the effects of maternal depressive symptoms on child development are attenuated. The absence of significant moderation in this study does not preclude this possibility, as the low prevalence of severe symptoms in this sample may have constrained our ability to detect meaningful moderation effects.

This null finding contrasts with evidence from previous literature identifying household composition as a potential buffer against the negative effects of maternal depressive symptoms (70, 71). Neither the number of young siblings nor the number of adults in the household significantly modified the association between maternal depressive symptoms and child development. However, as a covariate in the adjusted models, having two or more young siblings was negatively associated with child development in infancy but not in toddlerhood. This pattern may reflect the particularly high caregiving demands of early infancy, when multiple young children in the household simultaneously require intensive care and attention, potentially reducing the quality and frequency of one-to-one interactions available to the youngest child. As children grow into toddlerhood and develop greater autonomy, sibling presence may become less competing and more enriching, as older siblings can serve as social partners and models for learning (72). Extended family living arrangements are common in this setting, where caregiving responsibilities are often shared across multiple household members (46). Living in households with three or more adults showed a tendency toward lower child development scores at toddlerhood, a pattern that, while not statistically significant, may reflect two related dynamics. First, in this rural Andean context, multigenerational living arrangements may be driven by economic necessity rather than caregiving capacity, meaning that larger adult households may signal household vulnerability rather than additional resources for child development (20). Consistent with this interpretation, mothers with depressive symptoms in this sample were more likely to live in households with three or more adults, suggesting that extended family co-residence may not function as a protective buffer against maternal depressive symptoms themselves. Second, even when extended family support is present, qualitative evidence from this same context suggests such family support tends to prioritise addressing physical needs and household tasks over early stimulation activities, and that caregivers frequently struggle to find dedicated time for child-directed play and learning amid competing household demands (46). These findings highlight an important distinction: while distributed caregiving arrangements may buffer children from the effects of maternal depressive symptoms at the dyadic level, the mere presence of additional household members does not guarantee the quality or quantity of stimulating interactions that children need for healthy development. Together, these findings suggest that structural indicators of household composition alone are insufficient to capture the caregiving dynamics that shape early child development in this context.

The cultural context in which caregiving takes place may further shape both how maternal depressive symptoms is expressed and how it influences child-directed interactions. In the Andean region, depressive symptoms are often expressed through somatic complaints or socially mediated emotional expressions (73), which may not fully align with standardised questionnaire items. As a self-report measure, the DASS-21 captures mothers’ perceived experiences of depressive symptomatology rather than providing a clinical diagnosis (50) and responses can be influenced by cultural norms, social desirability, and individual perceptions of mental health (74, 75). These cultural influences may mean that the DASS-21 captures only a partial picture of maternal depressive symptoms in this setting. Such distress, in Andean communities, is often understood and expressed through relational and collective frameworks, where family networks play a central role in how it is both experienced and managed, patterns that standardised instruments designed in high-income contexts may not fully capture.

### Strengths, Limitations & Future Directions

This study has several strengths. First, it leverages a relatively large and well-characterised sample of mother–child dyads from an Andean setting, with data collected at two key developmental stages: infancy and toddlerhood. Furthermore, the inclusion of multiple contextual variables, including demographic, household, social, and caregiving factors, strengthens the interpretation of our findings and helps situate them within the broader environmental and cultural context.

However, we acknowledge the following limitations. First, the observational analytic approach precludes causal inference, and causal effects of maternal depressive symptoms on child development cannot be established from these data alone. Longitudinal designs with frequent measurement points, or quasi-experimental or experimental approaches would be better suited to establish temporal ordering and test causal pathways (76). Second, maternal depressive symptoms were assessed using a self-report instrument administered by interviewers which may be influenced by social desirability, cultural norms, and individual perceptions. Third, although the sample is representative of a specific rural Andean population, the analytic sample differed from the original RCT sample in age distribution and representation across study arms, which may limit the generalisability of findings. Fourth, the restricted range of depressive symptom scores in our sample, with most of the mothers reporting few or no symptoms, may have limited statistical power to detect moderation effects even if such were present. Finally, household composition was measured only at infancy and may not fully capture changes in family structure across the entire follow-up period to toddlerhood.

Future research should explore more detailed assessments of caregiving behaviours and family dynamics, including the quality and intensity of caregiving interactions across different household members. Specifically, understanding who provides stimulation activities to the child — and whether this changes when mothers experience depressive symptoms, would help clarify the mechanisms underlying the patterns observed in this study. Examining associations by specific domains of child development, such as language, motor, and socioemotional development, may also shed light on whether maternal depressive symptoms differentially affect developmental areas. Longer follow-up periods may help capture delayed or cumulative effects of maternal depressive symptoms on child outcomes. Additionally, examining culturally specific caregiving practices, household stability, and the distribution of caregiving responsibilities can provide insight into the compensatory mechanisms that may buffer children from the effects of maternal depressive symptoms in Andean and other LMIC contexts.

## Conclusion

This study examined the association between maternal depressive symptoms and early childhood development in a rural Andean population, finding no significant association at infancy or toddlerhood. Household composition did not moderate this association. Stimulation activities received by the child showed the strongest independent association with child development at both timepoints, regardless of maternal depressive symptom status, pointing to children’s exposure to stimulating interactions as a key correlate of early development in this context. Whether this reflects caregiving distributed across household and community networks warrants further investigation. The absence of a detectable association between maternal depressive symptoms and child development does not reduce the importance of supporting and priority of maternal mental health during the early years of parenting.

## Supporting information

See Supplementary Materials

## Author Contributions

MA contributed to Conceptualization, Formal Analysis, Investigation, Methodology, and Writing-Original Draft. KH contributed to Data Curation and Writing – Review & Editing. LJ contributed to Data Curation, Conceptualization, Writing – Review & Editing, and Supervision. GF, SH, DM, and DCM contributed to Conceptualization, Supervision, and Writing – Review & Editing. LA, SFH, AC, and MCG contributed to Writing – Review & Editing.

## Acknowledgments

We are grateful to the data collectors’ workers for their invaluable assistance in data collection and to the families who generously participated in the project. This project was funded by the Basel Research Centre for Child Health (BRCCH) through the Multi-Investigator Program. The funder played no role in the design, analysis or reporting of the results.

## Ethical considerations

This study was approved by the Institutional Review Boards of Universidad Peruana Cayetano Heredi (SIDISI: 214395) and Ethikkommission Nordwest- und Zentralschweiz (AO 2024-00100). Written informed consent was obtained from all participating families prior to data collection. The study was conducted in accordance with the Declaration of Helsinki and relevant local regulations to ensure participant confidentiality and safety.

## Data Availability Statement

The data that support the findings of this study are available from the corresponding author, upon reasonable request.

