## Supplementary material for "“The Role of Maternal Mental Health in Early Development: Evidence from High Altitude Peru”": See Supplementary Materials

**Summary**

This document provides supplementary materials for the manuscript examining the association between maternal depressive symptoms and early childhood development in 769 mother–child dyads from a cluster-randomised controlled trial conducted in the Cajamarca region of Peru. Maternal depressive symptoms were assessed during infancy (mean age 5.2 months) using the DASS-21 depression subscale. Child development was assessed concurrently during infancy using the CREDI Long Form and prospectively during toddlerhood (mean age 28.7 months) using the GSED Long Form. Associations were examined using linear regression models with cluster-robust standard errors, adjusting for demographic, household, social, and caregiving covariates.

The supplementary materials include descriptive statistics, full sequential model results for both timepoints, moderation analyses by household composition, and sensitivity analyses examining non-linear associations and moderation by intervention arm.

### 1. Descriptive Statistics

#### 1.1 Internal Consistency: DASS-21-Depression Subscale

The DASS-21 depression subscale demonstrated high consistency in this sample (α = 0.836, 95% CI [0.818, 0.854]), indicating that the seven items reliably measure depressive symptomatology.

**Table S1. Internal consistency: DASS-21 Depression Subscale**

| Statistic | Value |
|---|---|
| Cronbach's α | 0.836 |
| 95% CI | [0.818, 0.854] |
| Average inter-item correlation | 0.440 |
| Number of items | 7 |

#### 1.2. Baseline Characteristics by questionnaire completion

Table S2. Baseline characteristics by DASS-21 completion status

| | | DASS completion status | | |
|---|---|---|---|---|
| **Characteristic** | **N** | **DASS complete** N = 887[1] | **DASS missing** N = 1,815[1] | **p-value**[2] |
| **Child and maternal demographics** | | | | |
| **Child age (months)** | 2,699 | 5.10 (1.89) | 4.98 (1.96) | **0.047** |
| **Child sex** | 2,699 | | | 0.154 |
| Male | | 464 (52%) | 895 (49%) | |
| Female | | 423 (48%) | 917 (51%) | |
| **Mother's age** | 2,655 | | | 0.293 |
| Under 20 | | 115 (13%) | 199 (11%) | |
| 20-30 | | 450 (52%) | 955 (54%) | |
| 31 and older | | 308 (35%) | 628 (35%) | |
| **Mother's education** | 2,651 | | | 0.367 |
| None or Foundational | | 125 (14%) | 269 (15%) | |
| Intermediate | | 288 (33%) | 627 (35%) | |
| Higher | | 458 (53%) | 884 (50%) | |
| **Household characteristics** | | | | |
| **Place of residence** | 2,702 | | | 0.856 |
| Rural | | 551 (62%) | 1,134 (62%) | |
| Urban | | 336 (38%) | 681 (38%) | |

| | | DASS completion status | | |
|---|---|---|---|---|
| **Characteristic** | **N** | **DASS complete** N = 887[1] | **DASS missing** N = 1,815[1] | **p-value**[2] |
| **Household overcrowding** | 2,691 | | | 0.096 |
| No | | 541 (61%) | 1,171 (65%) | |
| Yes | | 340 (39%) | 639 (35%) | |
| **Socioeconomic index** | 2,689 | -0.03 (1.05) | 0.02 (0.97) | 0.100 |
| **Intervention arm** | **2,702** | | | **<0.001** |
| Control | | 293 (33%) | 671 (37%) | |
| Home visit intervention | | 139 (16%) | 178 (9.8%) | |
| Digital intervention | | 350 (39%) | 831 (46%) | |
| Cuna Mas | | 105 (12%) | 135 (7.4%) | |

[1]n (%); Mean (SD). DASS missing = participants who did not complete the depression scale at baseline. p-values: chi-squared for categorical, Wilcoxon for continuous.

[2]Wilcoxon rank sum test; Pearson's Chi-squared test

Dyads with complete DASS-21 data were broadly similar to those with missing data across most demographic and household characteristics. The statistically significant differences were observed in child age at baseline, children with complete data were slightly older (mean 5.10 vs. 4.98 months, p = 0.047), and in intervention arm distribution (p < 0.001), with a lower proportion of complete data in the digital intervention arm and control arms compared to the home visit and Cuna Más arms. No significant differences were observed in maternal age, maternal education, place of residence, household overcrowding, or socioeconomic status. These findings suggest that missing data on DASS-21 were not systematically related to most sociodemographic characteristics, though the differential completion by intervention arm should be noted when interpreting findings.

### 1.3. Baseline characteristics by intervention arm

Table S3. Baseline characteristics by intervention arm

| **Characteristic** | **Control** N = 267[1] | **Home visit intervention** N = 130[1] | **Digital intervention** N = 291[1] | **Cuna Mas** N = 81[1] | **p-value**[2] |
|---|---|---|---|---|---|
| **Demographics** | | | | | |
| **Child sex** | | | | | 0.303 |
| Male | 128 (48%) | 66 (51%) | 159 (55%) | 46 (57%) | |
| Female | 139 (52%) | 64 (49%) | 132 (45%) | 35 (43%) | |
| **Mother's age** | | | | | 0.612 |
| Under 20 | 35 (13%) | 13 (10%) | 39 (14%) | 8 (10%) | |

| Characteristic | Control N = 267[1] | Home visit intervention N = 130[1] | Digital intervention N = 291[1] | Cuna Mas N = 81[1] | p-value[2] |
|---|---|---|---|---|---|
| 20-30 | 133 (50%) | 64 (50%) | 152 (54%) | 38 (48%) | |
| 31 and older | 96 (36%) | 52 (40%) | 93 (33%) | 34 (43%) | |
| **Mother's education level** | | | | | **<0.001** |
| None or Foundational | 39 (15%) | 24 (19%) | 24 (8.5%) | 21 (26%) | |
| Intermediate | 92 (35%) | 37 (29%) | 85 (30%) | 34 (43%) | |
| Higher | 133 (50%) | 68 (53%) | 175 (62%) | 25 (31%) | |
| **Planned pregnancy** | | | | | **0.045** |
| No | 130 (49%) | 75 (58%) | 169 (59%) | 38 (48%) | |
| Yes | 137 (51%) | 54 (42%) | 118 (41%) | 42 (53%) | |
| **Household characteristics** | | | | | |
| **Place of residence** | | | | | **<0.001** |
| Rural | 196 (73%) | 76 (58%) | 137 (47%) | 75 (93%) | |
| Urban | 71 (27%) | 54 (42%) | 154 (53%) | 6 (7.4%) | |
| **Household overcrowding** | | | | | 0.948 |
| No | 163 (61%) | 83 (64%) | 180 (63%) | 49 (61%) | |
| Yes | 104 (39%) | 47 (36%) | 107 (37%) | 31 (39%) | |
| **Socioeconomic index** | -0.21 (0.99) | 0.09 (1.07) | 0.24 (1.05) | -0.67 (0.80) | **<0.001** |
| **Father is head of household** | | | | | 0.798 |
| No | 26 (11%) | 14 (12%) | 30 (12%) | 6 (8.1%) | |
| Yes | 207 (89%) | 101 (88%) | 217 (88%) | 68 (92%) | |
| **Household composition** | | | | | |
| **Sibling under 6 years** | | | | | 0.023 |
| 0 | 175 (66%) | 99 (76%) | 223 (77%) | 49 (60%) | |
| 1 | 80 (30%) | 26 (20%) | 61 (21%) | 28 (35%) | |
| 2+ | 12 (4.5%) | 5 (3.8%) | 7 (2.4%) | 4 (4.9%) | |

| Characteristic | Control<br>N = 267[1] | Home visit intervention<br>N = 130[1] | Digital intervention<br>N = 291[1] | Cuna Mas<br>N = 81[1] | p-value[2] |
|---|---|---|---|---|---|
| **Children 6-12 years** | 0.61 (0.75) | 0.67 (0.86) | 0.61 (0.78) | 0.88 (0.84) | **0.033** |
| **Children 13-18 years** | 1.46 (0.79) | 1.43 (0.65) | 1.36 (0.67) | 1.36 (0.66) | 0.225 |
| **Adults in the household** | | | | | 0.344 |
| 1-2 | 192 (72%) | 85 (65%) | 203 (70%) | 62 (77%) | |
| 3+ | 75 (28%) | 45 (35%) | 86 (30%) | 19 (23%) | |
| **Social and stimulation factors** | | | | | |
| **Stimulation activities** | 3.32 (1.40) | 3.40 (1.26) | 3.63 (1.39) | 2.95 (1.12) | **<0.001** |
| **Social support** | 14 (9-19) | 12 (8-20) | 15 (10-19) | 12 (8-19) | 0.388 |
| **Social assistance programme participation** | | | | | **<0.001** |
| No | 164 (62%) | 87 (67%) | 189 (66%) | 9 (11%) | |
| Yes | 102 (38%) | 43 (33%) | 98 (34%) | 71 (89%) | |
| **Child development outcomes** | | | | | |
| **Age-adjusted development score (Infancy)** | -0.02 (0.59) | -0.08 (0.58) | 0.10 (0.57) | -0.24 (0.58) | **<0.001** |
| **Age-adjusted development score (Toddlerhood)** | -0.3 (4.3) | -0.2 (5.7) | 0.4 (5.2) | -0.2 (5.3) | **0.035** |

[1]n (%); Mean (SD)

[2]Pearson's Chi-squared test; Kruskal-Wallis rank sum test; Fisher's exact test

Baseline characteristics were largely balanced across the four randomised arms (control, home visit intervention, digital intervention, and Cuna Mas), with some expected variation. The Cuna Más arm differed from the other arms across characteristics, including a higher proportion of rural residents (93%), lower socioeconomic status, lower maternal education, and higher social assistance programme participation (89%), reflecting the programme's targeting of households in poverty and extreme poverty. Given these systematic differences, intervention arm is included as a covariate in all toddlerhood models, and its potential moderating role is examined as a sensitivity analysis (Table S5).

### 2. Sensitivity Analysis

#### 2.1. Non-linear association between maternal depressive symptoms and child development

Neither the quadratic term nor the natural cubic spline specification improved model fit over the linear model at either timepoint. For the infancy model, the quadratic term was non-significant (β

= −0.000, p = 0.391) and the AIC increased slightly (1,651.2 vs. 1,652.7), supporting retention of the linear specification. For the toddlerhood model, the quadratic term was similarly non-significant (β = −0.004, p = 0.227) and the AIC showed no meaningful improvement (4,265.5 vs. 4,264.8). These findings suggest that the association between maternal depressive symptoms and child development is adequately captured by a linear specification at both timepoints.

Table S4. Sensitivity analysis: non-linear association between maternal depressive symptoms and child development

| Outcome | Term | β | SE | p-value | AIC (linear) | AIC (quadratic) |
|---|---|---|---|---|---|---|
| Development score (Infancy) | Depression score (linear) | 0.0052 | 0.0083 | 0.5259 | 1,651.2 | 1,652.7 |
| | Depression score² | -0.0003 | 0.0003 | 0.3913 | | |
| Development score (Infancy) (Toddlerhood) | Depression score (linear) | 0.0825 | 0.0677 | 0.2234 | 4,265.5 | 4,264.8 |
| | Depression score² | -0.0038 | 0.0031 | 0.2272 | | |

Non-significant quadratic term (p > 0.05) with no AIC improvement supports retention of the linear specification. Models adjusted for full covariate set.

### 2.2. Moderation by intervention arm at toddlerhood

Intervention arm did not significantly moderate the association between maternal depressive symptoms and child development at toddlerhood for the home visit or digital intervention arms. A significant interaction was observed for the Cuna Más arm (β = −0.12, 95% CI [−0.19, −0.05], p = 0.001), suggesting that the association between maternal depressive symptoms and child development differed in this arm compared to the control group.

Table S5. Sensitivity analysis: moderation of the association between maternal depressive symptoms and child development by intervention arm at toddlerhood

| Characteristic | Beta | 95% CI | p-value |
|---|---|---|---|
| Maternal depressive symptoms | 0.05 | -0.01, 0.11 | 0.079 |
| Intervention arm | | | |
| Control | Ref. | | |
| Maternal depressive symptoms x Home visit intervention | -0.17 | -1.6, 1.2 | 0.812 |
| Maternal depressive symptoms x Digital intervention | 0.50 | -0.34, 1.3 | 0.243 |
| Maternal depressive symptoms x Cuna Mas | 0.21 | -0.54, 0.97 | 0.582 |
| Depression x Intervention arm | | | |
| Maternal depressive symptoms x Home visit intervention | -0.11 | -0.24, 0.01 | 0.068 |
| Maternal depressive symptoms x Digital intervention | -0.10 | -0.25, 0.04 | 0.175 |
| Maternal depressive symptoms x Cuna Mas | -0.12 | -0.19, -0.05 | **0.001** |

| Characteristic | Beta | 95% CI | p-value |
|---|---|---|---|

Abbreviation: CI = Confidence Interval

### 3. Estimated Association between Maternal Depressive Symptoms and Child Development in Infancy

#### 3.1 Sequential Models

The depression coefficient remained consistently near zero and non-significant across all four sequential models, indicating that the null concurrent association between maternal depressive symptoms and child development in infancy is robust to progressive covariate adjustment. Maternal education (higher vs. none/foundational) was positively associated with child development in earlier models but attenuated to non-significance in the fully adjusted model ($\beta = 0.15$, 95% CI [−0.06, 0.35], $p = 0.166$), suggesting this effect may operate partly through stimulation practices and social support. The presence of two or more siblings under 6 years in the household was negatively associated with child development and remained significant in the fully adjusted model ($\beta = -0.42$, 95% CI [−0.81, −0.02], $p = 0.040$), consistent with a caregiving burden effect. Stimulation activities showed the strongest independent association with child development ($\beta = 0.12$, 95% CI [0.08, 0.17], $p < 0.001$).

Table S6. Full model results for the association between maternal depressive symptoms and child development in infancy

| | Model 1 (Unadjusted) | | | Model 2 (Demographics) | | | Model 3 Household characteristics | | | Model 4 (Household composition and social & stimulation factors) | | |
|---|---|---|---|---|---|---|---|---|---|---|---|---|
| **Characteristic** | **Beta** | **95% CI** | **p-value** | **Beta** | **95% CI** | **p-value** | **Beta** | **95% CI** | **p-value** | **Beta** | **95% CI** | **p-value** |
| **Exposure** | | | | | | | | | | | | |
| Maternal depressive symptoms (Infancy) | 0.00 | -0.01, 0.01 | 0.601 | 0.00 | -0.01, 0.01 | 0.613 | 0.00 | -0.01, 0.01 | 0.573 | 0.00 | -0.01, 0.01 | 0.753 |
| **Demographics** | | | | | | | | | | | | |
| Child age (months) | 0.72 | 0.68, 0.75 | **<0.001** | 0.72 | 0.68, 0.75 | **<0.001** | 0.70 | 0.67, 0.74 | **<0.001** | 0.69 | 0.66, 0.73 | **<0.001** |
| Child sex | | | | | | | | | | | | |
| Males | | | | Ref. | | | | | | | | |
| Female | | | | -0.04 | -0.17, 0.09 | 0.523 | -0.02 | -0.15, 0.11 | 0.752 | -0.01 | -0.13, 0.11 | 0.875 |
| Mother age | | | | | | | | | | | | |
| Under 20 | | | | Ref. | | | | | | | | |
| 20-30 | | | | -0.07 | -0.26, 0.12 | 0.454 | -0.03 | -0.23, 0.16 | 0.733 | -0.06 | -0.26, 0.14 | 0.566 |
| 31 and older | | | | -0.11 | -0.32, 0.10 | 0.304 | -0.10 | -0.32, 0.11 | 0.353 | -0.06 | -0.28, 0.17 | 0.634 |
| Mother education | | | | | | | | | | | | |
| None or Foundational | | | | Ref. | | | | | | | | |
| Intermediate | | | | 0.12 | -0.06, 0.29 | 0.189 | 0.08 | -0.09, 0.24 | 0.361 | 0.03 | -0.14, 0.21 | 0.724 |

| | Model 1 (Unadjusted) | | | Model 2 (Demographics) | | | Model 3 Household characteristics | | | Model 4 (Household composition and social & stimulation factors) | | |
|---|---|---|---|---|---|---|---|---|---|---|---|---|
| **Characteristic** | **Beta** | **95% CI** | **p-value** | **Beta** | **95% CI** | **p-value** | **Beta** | **95% CI** | **p-value** | **Beta** | **95% CI** | **p-value** |
| Higher | | | | 0.39 | 0.23, 0.55 | **<0.001** | 0.23 | 0.05, 0.41 | **0.012** | 0.15 | -0.06, 0.35 | 0.166 |
| **Household characteristics** | | | | | | | | | | | | |
| Place of residence | | | | | | | | | | | | |
| Rural | | | | | | | Ref. | | | | | |
| Urban | | | | | | | 0.04 | -0.19, 0.27 | 0.723 | 0.03 | -0.16, 0.23 | 0.744 |
| Socioeconomic index | | | | | | | 0.09 | 0.01, 0.17 | **0.031** | 0.02 | -0.06, 0.09 | 0.627 |
| **Household composition** | | | | | | | | | | | | |
| Sibling under 6 years | | | | | | | | | | | | |
| 0 | | | | | | | Ref. | | | | | |
| 1 | | | | | | | -0.17 | -0.31, -0.04 | **0.012** | -0.11 | -0.25, 0.02 | 0.094 |
| 2+ | | | | | | | -0.55 | -0.92, -0.18 | **0.004** | -0.42 | -0.81, -0.02 | **0.040** |
| Children 6-12 years | | | | | | | -0.02 | -0.09, 0.06 | 0.689 | 0.00 | -0.08, 0.09 | 0.915 |
| Children 13-18 years | | | | | | | 0.04 | -0.05, 0.13 | 0.403 | 0.03 | -0.06, 0.12 | 0.546 |
| Adults in the household | | | | | | | | | | | | |
| 1-2 | | | | | | | Ref. | | | | | |

| | Model 1 (Unadjusted) | | | Model 2 (Demographics) | | | Model 3 Household characteristics | | | Model 4 (Household composition and social & stimulation factors) | | |
|---|---|---|---|---|---|---|---|---|---|---|---|---|
| **Characteristic** | **Beta** | **95% CI** | **p-value** | **Beta** | **95% CI** | **p-value** | **Beta** | **95% CI** | **p-value** | **Beta** | **95% CI** | **p-value** |
| 3+ | | | | | | | -0.06 | -0.18, 0.06 | 0.307 | -0.05 | -0.17, 0.07 | 0.453 |
| **Social and stimulation factors** | | | | | | | | | | | | |
| Stimulation activities (0-6) | | | | | | | | | | 0.12 | 0.08, 0.17 | **<0.001** |
| Social support score | | | | | | | | | | 0.01 | 0.00, 0.02 | **0.006** |
| Social assistance programme participation | | | | | | | | | | | | |
| No | | | | | | | | | | Ref. | | |
| Yes | | | | | | | | | | -0.14 | -0.29, 0.02 | 0.085 |

Abbreviation: CI = Confidence Interval

### 4. Estimated Association between Maternal Depressive Symptoms and Child Development in Toddlerhood

#### 4.1 Sequential Models

The depression coefficient remained consistently near zero and non-significant across all six sequential models (β = −0.01, p = 0.638–0.786), confirming that the null prospective association between maternal depressive symptoms in infancy and child development at toddlerhood is robust to progressive covariate adjustment. Maternal education (higher vs. none/foundational) was positively associated with child development in the demographics model (β = 1.1, 95% CI [0.06, 2.2], p = 0.038) but attenuated to non-significance after adding household and social factors (β = 0.41, 95% CI [−0.89, 1.7], p = 0.532), suggesting this effect may operate through socioeconomic and stimulation pathways. Socioeconomic status showed a positive association with child development that reached significance in the fully adjusted model (β = 0.46, 95% CI [0.01, 0.91], p = 0.046). Stimulation activities showed the strongest independent association with child development and remained stable across models (β = 0.38–0.39, p = 0.008). The presence of three or more adults in the household showed a tendency toward lower child development scores, though this did not reach conventional statistical significance in the fully adjusted model (β = −0.94, 95% CI [−1.9, 0.00], p = 0.051). Intervention arm coefficients were non-significant, supporting its inclusion as a covariate rather than a moderator.

Table S7. Full model results for the association between maternal depressive symptoms and child development in toddlerhood

| | M1 Unadjusted | | | M2 Demographics | | | M3 Household characteristics | | | M4 Household composition | | | M5 Socio and stimulation factors | | | M6 Intervention arm | | |
|---|---|---|---|---|---|---|---|---|---|---|---|---|---|---|---|---|---|---|
| **Characteristic** | **Beta** | **95% CI** | **p-value** | **Beta** | **95% CI** | **p-value** | **Beta** | **95% CI** | **p-value** | **Beta** | **95% CI** | **p-value** | **Beta** | **95% CI** | **p-value** | **Beta** | **95% CI** | **p-value** |
| **Exposure** | | | | | | | | | | | | | | | | | | |
| Maternal depressive symptoms (Infancy) | -0.01 | -0.07, 0.04 | 0.638 | -0.01 | -0.07, 0.04 | 0.646 | -0.01 | -0.07, 0.04 | 0.642 | -0.01 | -0.07, 0.05 | 0.786 | -0.01 | -0.07, 0.05 | 0.694 | -0.01 | -0.07, 0.05 | 0.728 |
| **Demographics** | | | | | | | | | | | | | | | | | | |
| Child age (days) | 0.03 | 0.02, 0.03 | <0.001 | 0.03 | 0.02, 0.03 | <0.001 | 0.03 | 0.02, 0.03 | <0.001 | 0.03 | 0.02, 0.03 | <0.001 | 0.03 | 0.02, 0.03 | <0.001 | 0.03 | 0.02, 0.03 | <0.001 |
| Child sex | | | | | | | | | | | | | | | | | | |
| Male | | | | Ref. | | | | | | | | | | | | | | |
| Female | | | | 0.31 | -0.41, 1.0 | 0.403 | 0.37 | -0.34, 1.1 | 0.309 | 0.39 | -0.31, 1.1 | 0.273 | 0.33 | -0.35, 1.0 | 0.340 | 0.37 | -0.31, 1.0 | 0.291 |
| Mother age | | | | | | | | | | | | | | | | | | |

| | M1 Unadjusted | | | M2 Demographics | | | M3 Household characteristics | | | M4 Household composition | | | M5 Socio and stimulation factors | | | M6 Intervention arm | | |
|---|---|---|---|---|---|---|---|---|---|---|---|---|---|---|---|---|---|---|
| **Characteristic** | **Beta** | **95% CI** | **p-value** | **Beta** | **95% CI** | **p-value** | **Beta** | **95% CI** | **p-value** | **Beta** | **95% CI** | **p-value** | **Beta** | **95% CI** | **p-value** | **Beta** | **95% CI** | **p-value** |
| Under 20 | | | | Ref. | | | | | | | | | | | | | | |
| 20-30 | | | | 0.06 | -1.0, 1.2 | 0.917 | -0.08 | -1.2, 1.1 | 0.895 | -0.19 | -1.4, 1.0 | 0.760 | -0.19 | -1.4, 1.0 | 0.760 | -0.16 | -1.4, 1.1 | 0.795 |
| 31 and older | | | | -0.10 | -1.4, 1.2 | 0.881 | -0.20 | -1.5, 1.1 | 0.760 | -0.33 | -1.7, 1.0 | 0.632 | -0.59 | -2.1, 0.88 | 0.433 | -0.56 | -2.0, 0.91 | 0.455 |
| Mother education | | | | | | | | | | | | | | | | | | |
| None or Foundational | | | | Ref. | | | | | | | | | | | | | | |
| Intermediate | | | | -0.10 | -1.1, 0.85 | 0.830 | -0.23 | -1.2, 0.73 | 0.633 | -0.25 | -1.2, 0.73 | 0.619 | -0.27 | -1.3, 0.78 | 0.610 | -0.33 | -1.4, 0.72 | 0.536 |
| Higher | | | | 1.1 | 0.06, 2.2 | **0.038** | 0.73 | -0.48, 1.9 | 0.238 | 0.73 | -0.48, 2.0 | 0.236 | 0.46 | -0.80, 1.7 | 0.471 | 0.41 | -0.89, 1.7 | 0.532 |
| **Household characteristics** | | | | | | | | | | | | | | | | | | |
| Place of residence | | | | | | | | | | | | | | | | | | |
| Rural | | | | | | | Ref. | | | | | | | | | | | |
| Urban | | | | | | | -0.12 | -1.0, 0.79 | 0.799 | -0.17 | -1.0, 0.71 | 0.712 | -0.45 | -1.4, 0.51 | 0.359 | -0.54 | -1.5, 0.38 | 0.250 |
| Socioeconomic index | | | | | | | 0.38 | -0.13, 0.88 | 0.143 | 0.38 | -0.12, 0.88 | 0.133 | 0.46 | 0.02, 0.90 | **0.042** | 0.46 | 0.01, 0.91 | **0.046** |
| **Household composition** | | | | | | | | | | | | | | | | | | |
| Sibling under 6 years | | | | | | | | | | | | | | | | | | |
| 0 | | | | | | | | | | Ref. | | | | | | | | |
| 1 | | | | | | | | | | -0.42 | -1.3, 0.42 | 0.329 | -0.45 | -1.4, 0.47 | 0.335 | -0.45 | -1.4, 0.47 | 0.337 |
| 2+ | | | | | | | | | | -1.2 | -3.1, 0.70 | 0.216 | -1.2 | -3.2, 0.67 | 0.202 | -1.2 | -3.1, 0.66 | 0.202 |

| | M1 Unadjusted | | | M2 Demographics | | | M3 Household characteristics | | | M4 Household composition | | | M5 Socio and stimulation factors | | | M6 Intervention arm | | |
|---|---|---|---|---|---|---|---|---|---|---|---|---|---|---|---|---|---|---|
| **Characteristic** | **Beta** | **95% CI** | **p-value** | **Beta** | **95% CI** | **p-value** | **Beta** | **95% CI** | **p-value** | **Beta** | **95% CI** | **p-value** | **Beta** | **95% CI** | **p-value** | **Beta** | **95% CI** | **p-value** |
| Children 6-12 years | | | | | | | | | | 0.16 | -0.30, 0.62 | 0.500 | 0.31 | -0.14, 0.75 | 0.178 | 0.29 | -0.16, 0.75 | 0.206 |
| Children 13-18 years | | | | | | | | | | -0.16 | -0.67, 0.36 | 0.551 | -0.02 | -0.51, 0.48 | 0.951 | -0.01 | -0.52, 0.49 | 0.957 |
| Adults at the household | | | | | | | | | | | | | | | | | | |
| 1-2 | | | | | | | | | | Ref. | | | | | | | | |
| 3+ | | | | | | | | | | -0.62 | -1.6, 0.33 | 0.199 | -0.96 | -1.9, -0.04 | **0.041** | -0.94 | -1.9, 0.00 | 0.051 |
| **Social and stimulation factors** | | | | | | | | | | | | | | | | | | |
| Stimulation activities | | | | | | | | | | | | | 0.39 | 0.10, 0.67 | **0.008** | 0.38 | 0.10, 0.67 | **0.008** |
| Social support | | | | | | | | | | | | | -0.03 | -0.09, 0.03 | 0.323 | -0.03 | -0.09, 0.03 | 0.316 |
| Social assistance programme participation | | | | | | | | | | | | | | | | | | |
| No | | | | | | | | | | | | | Ref. | | | | | |
| Yes | | | | | | | | | | | | | -0.01 | -0.64, 0.63 | 0.978 | -0.08 | -0.79, 0.62 | 0.816 |
| **Intervention arm** | | | | | | | | | | | | | | | | | | |
| Control | | | | | | | | | | | | | | | | Ref. | | |
| Home visit intervention | | | | | | | | | | | | | | | | -0.20 | -1.6, 1.1 | 0.767 |
| Digital intervention | | | | | | | | | | | | | | | | 0.45 | -0.39, 1.3 | 0.291 |

| | M1<br>Unadjusted | | | M2<br>Demographics | | | M3<br>Household characteristics | | | M4<br>Household composition | | | M5<br>Socio and stimulation factors | | | M6<br>Intervention arm | | |
|---|---|---|---|---|---|---|---|---|---|---|---|---|---|---|---|---|---|---|
| **Characteristic** | **Beta** | **95% CI** | **p-value** | **Beta** | **95% CI** | **p-value** | **Beta** | **95% CI** | **p-value** | **Beta** | **95% CI** | **p-value** | **Beta** | **95% CI** | **p-value** | **Beta** | **95% CI** | **p-value** |
| Cuna Mas | | | | | | | | | | | | | | | | 0.30 | -0.49, 1.1 | 0.454 |

Abbreviation: CI = Confidence Interval

### 5. Moderation of the association between maternal depressive symptoms and child development

#### 5.1 Interaction plots in Infancy

Figures S1 and S2 display interaction plots for the moderation analyses in infancy. In both figures, the regression lines across groups are largely parallel and overlapping, with wide and overlapping confidence intervals, consistent with the absence of statistically significant moderation effects.

Figure S1 shows that child development scores in infancy were slightly lower in households with two or more siblings under 6 years (green line), reflecting the main effect observed in the adjusted models. However, the slopes across the three sibling groups are nearly identical, indicating that the association between maternal depressive symptoms and child development did not differ by number of young siblings.

Figure S2 shows similarly parallel slopes for households with 1-2 versus 3 or more adults. Although children in households with 3 or more adults showed marginally lower development scores at lower depression levels, this pattern did not reach statistical significance, and the confidence intervals overlap substantially across the full range of depression scores.

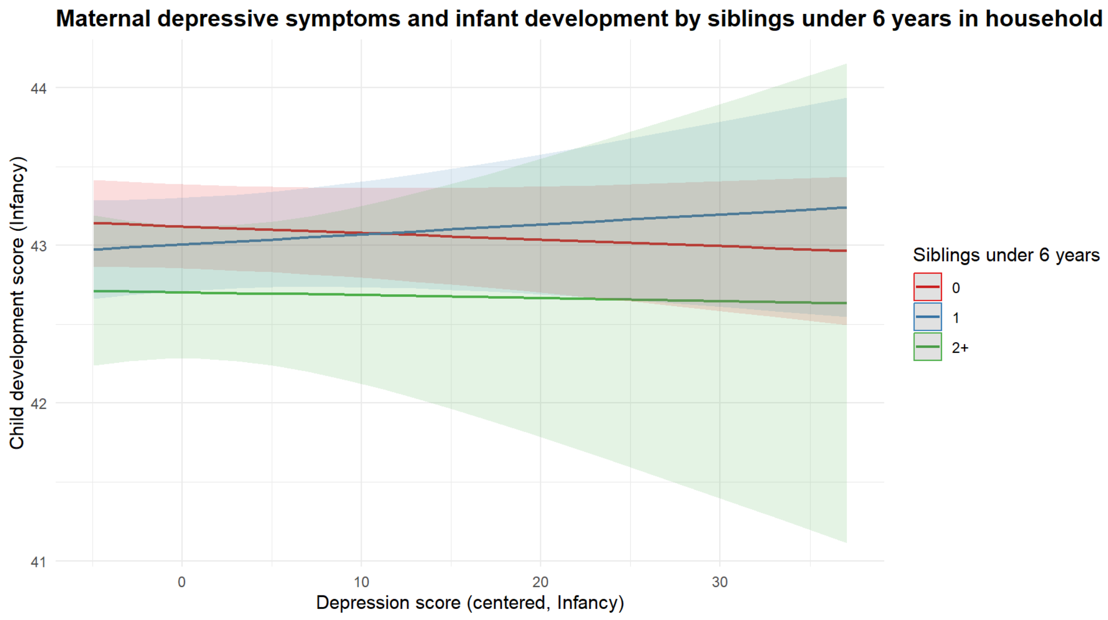

*Figure S1. Maternal depressive symptoms and child development by siblings under 6 years in household (adjusted, infancy).*

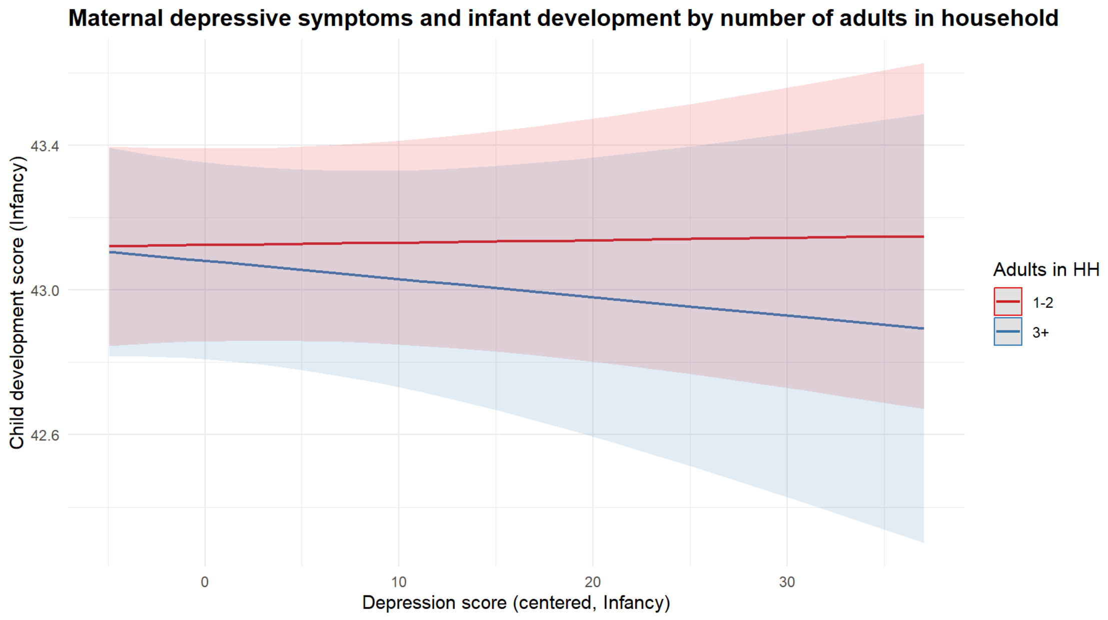

*Figure S2. Maternal depressive symptoms and child development by number of adults in household (adjusted, infancy).*

### 5.2 Interaction plots in toddlerhood

Figures S3 and S4 display interaction plots for the moderation analyses in toddlerhood. As in infancy, the regression lines across groups are largely parallel with wide and overlapping confidence intervals, consistent with the absence of statistically significant moderation effects.

Figure S3 shows that child development scores in toddlerhood were broadly similar across sibling groups, with overlapping confidence intervals across the full range of depression scores. The slopes across the three groups are nearly identical, indicating that the association between maternal depressive symptoms and child development did not differ by number of young siblings in the household.

Figure S4 shows that children in households with 1–2 adults had marginally higher development scores than those in households with 3 or more adults across the full range of depression scores. However, the slopes for both groups are similarly flat, indicating no evidence that the number of adults in the household moderated the association between maternal depressive symptoms and child development at toddlerhood.

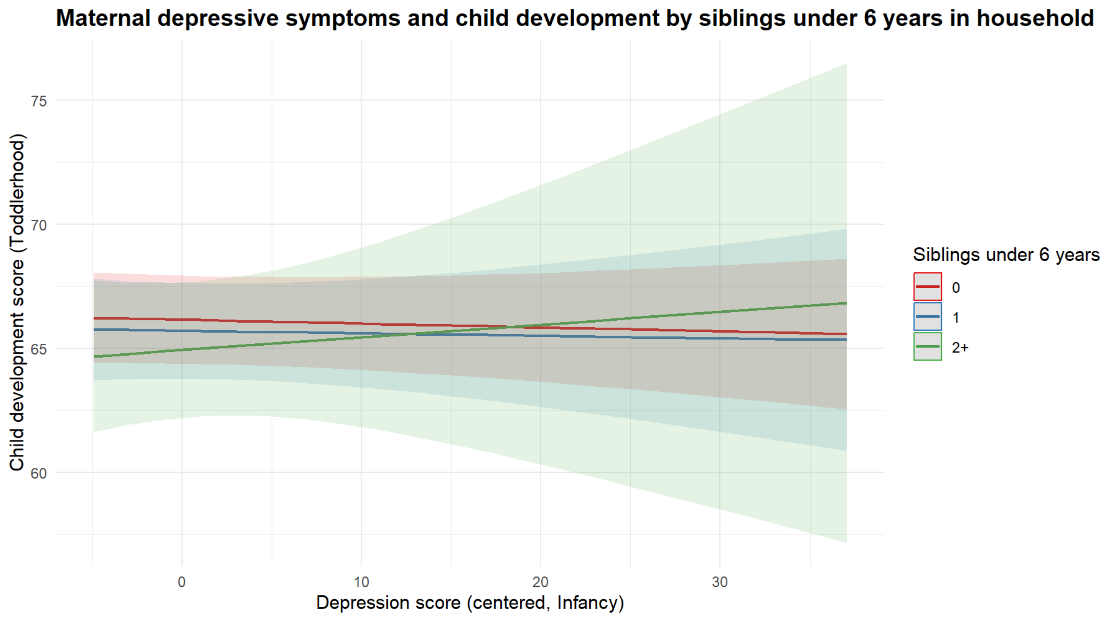

*Figure S3. Maternal depressive symptoms and child development by siblings under 6 years in household (adjusted, toddlerhood).*

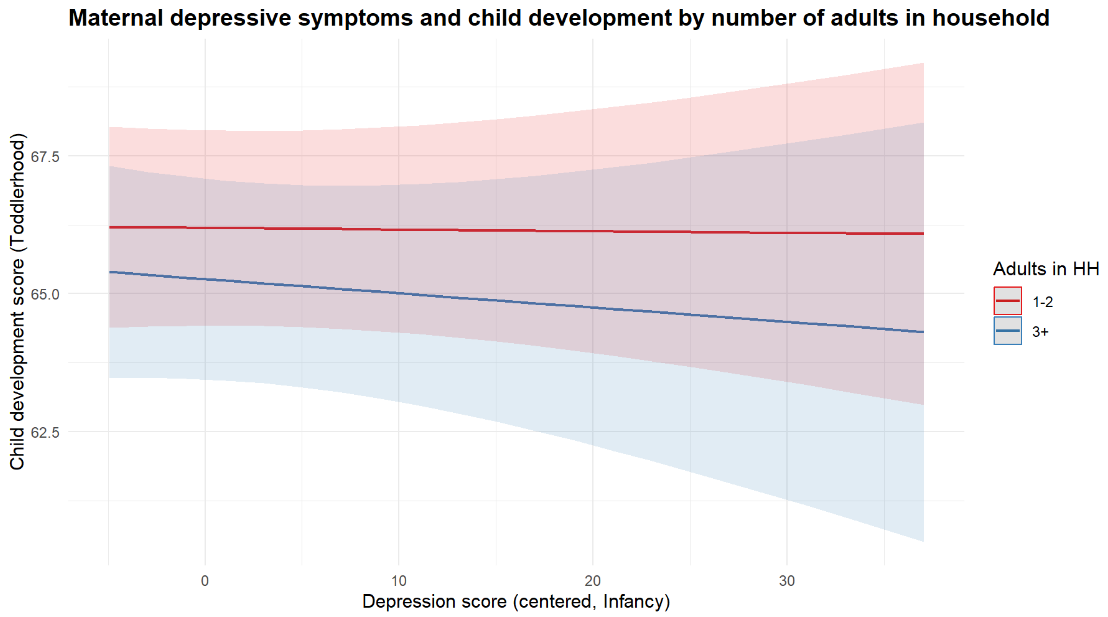

*Figure S4. Maternal depressive symptoms and child development by number of adults in household (adjusted, toddlerhood).*
